# Clinical utility of plasma Aβ42/40 ratio by LC-MS/MS in Alzheimer’s disease assessment

**DOI:** 10.1101/2023.12.12.23299878

**Authors:** Darren M. Weber, Steven W. Taylor, Robert J. Lagier, Jueun C. Kim, Scott M. Goldman, Nigel J. Clarke, David E. Vaillancourt, Ranjan Duara, Karen N. McFarland, Wei-en Wang, Todd E. Golde, Michael K. Racke

**Affiliations:** Quest Diagnostics Nichols Institute, San Juan Capistrano, CA USA; Department of Applied Physiology and Kinesiology, Fixel Institute for Neurological Disorders, and 1Florida ADRC, University of Florida, Gainesville, FL USA; Wien Center for Alzheimer’s Disease and Memory Disorders, Mount Sinai Medical Center, Miami Beach, FL USA; Department of Neurology, Center for Translational Research in Neurodegenerative Disease, 1Florida Alzheimer’s Disease Research Center (ADRC), University of Florida, Gainesville, FL USA; Department of Pharmacology and Chemical Biology and Department of Neurology Center for Neurodegenerative Disease, Goizueta Institute Emory Brain Health, Emory University, School of Medicine. Atlanta, GA USA

**Keywords:** Alzheimer’s disease, amyloid, biomarkers, blood biomarkers, LC-MS/MS, PET, prescreening

## Abstract

**INTRODUCTION:** Plasma Aβ42/40 ratio can be used to help predict amyloid PET status, but its clinical utility in Alzheimer’s disease (AD) assessment is unclear.

**METHODS:** Aβ42/40 ratio was measured by LC-MS/MS in 250 specimens with associated amyloid PET imaging, diagnosis, and demographic data, and 6,192 consecutive clinical specimens submitted for Aβ42/40 testing.

**RESULTS:** High diagnostic sensitivity and negative predictive value (NPV) for Aβ-PET positivity were observed, consistent with the clinical performance of other plasma LC-MS/MS assays, but with greater separation between Aβ42/40 values for individuals with positive vs negative Aβ-PET results. Assuming a moderate prevalence of Aβ-PET positivity, a cutpoint was identified with 99% NPV, which could help predict that AD is likely not the cause of patients’ cognitive impairment and help reduce PET evaluation by about 40%.

**DISCUSSION:** Using high-throughput plasma Aβ42/40 LC-MS/MS assays can help reduce PET evaluations in patients with low likelihood of AD pathology, allowing for cost savings.

**Highlights:**

- A new plasma LC-MS/MS assay for the Aβ42/Aβ40 ratio has clinical utility in AD assessment.
- Performance was assessed using specimens with a moderate prevalence of Aβ-PET positivity.
- Analysis was extrapolated to 6,192 consecutive clinical specimens submitted for ratio testing.
- Assay cutpoints were proposed to help suggest clinical management decisions.
- Based on the assay’s high NPV, costly PET evaluations may be avoided for many individuals.

**Research in Context:** *Systematic Review:* Aβ42/Aβ40 ratio data were analyzed from 250 ADRC cohort participants and 6,192 plasma specimens submitted for Aβ42/40 ratio testing by LC-MS/MS at Quest Diagnostics.

*Interpretation:* Based on its high NPV, the assay identifies individuals likely to have negative amyloid PET results. Higher discriminatory power and larger fold-changes between PET-positive and negative individuals were observed compared with previous studies. Our “real-world” data set, combined with known performance characteristics, allows us to suggest cutpoints and clinical decisions based on plasma Aβ42/40 ratios.

*Future Directions:* Longitudinal plasma specimens from individuals who convert from PET-negative to positive, or that transition from cognitively normal to MCI and AD, will improve understanding of the prognostic utility of the Aβ42/40 ratio. Using Aβ42/40 ratio alone or combined with other biomarkers, to follow patients with cognitive impairment may yield insights regarding their disease conditions and progression and who may benefit from disease-modifying therapeutics.

## 1 INTRODUCTION

Alzheimer’s disease (AD) pathophysiology is characterized by cognitive impairment and the accumulation of extracellular beta-amyloid (Aβ) plaques and intracellular neurofibrillary tangles composed of hyperphosphorylated tau protein in brain tissue.^1–3^ Because Aβ aggregation and deposition occurs 10-20 years prior to clinical presentation, plaque detection reflects an underlying pathophysiologic process at a prodromal or pre-clinical disease stage ^4–6^ and a potential opportunity for early intervention and treatment. In addition, ruling out AD can prompt investigation of non-AD causes of dementia and cognitive decline.^7^

Recently, the United States Food and Drug Administration (FDA) approved the first disease-modifying treatments for AD, using monoclonal antibodies targeting Aβ aggregates and removing them from the brain. Administering monoclonal antibodies that target Aβ slowed the rate of functional and cognitive decline among patients with AD in the mild cognitive impairment (MCI) or mild dementia stage. Presence of Aβ protein was assessed using positron emission tomography (PET).^8,9^ PET and measurement of cerebrospinal fluid (CSF) beta-amyloid 42 (Aβ42)^10,11^ are methods that have been used as entry criteria for clinical trials and/or as outcome measures for disease-modifying AD treatments.^12^

Blood-based biomarkers, including the plasma beta-amyloid ratio (Aβ42/40), may guide, complement, or be alternatives to PET and CSF testing. However, despite its lower cost and decreased invasive nature, routine clinical use of plasma Aβ42/40 testing has been suggested to have substantial challenges.^13^ Immunoassay platforms for measurement of Aβ42/40 have been evaluated for assay robustness, with small measurement or preanalytical errors leading to misclassification risks.^13^ Evaluation of liquid chromatography-tandem mass spectrometry (LC-MS/MS) platforms has been more limited,^13^ despite repeatedly demonstrating superior performance characteristics compared to immunoassays.^13–15^

We developed and validated a protein immunoprecipitation (IP)-LC-MS/MS assay for the detection of plasma Aβ40 and Aβ42 (Weber et al, submitted).^16^ Here we assessed the clinical performance of using the Aβ42/40 ratio to identify amyloid PET status in a well-characterized cohort from the 1Florida Alzheimer’s Disease Research Center (ADRC cohort). The cohort included age-matched Aβ PET-positive and Aβ PET-negative individuals characterized as AD, MCI, or healthy controls with *APOE* genotype inferred from apolipoprotein E (ApoE) isoform proteotype. Part of the assessment included examining the effect of imprecision and bias estimates on classifying associated risk for PET positivity.^13^ Based on these performance characteristics, we evaluated incorporating Aβ42/Aβ40 results into AD assessment using a limited data set from 6,192 specimens submitted for Aβ42/Aβ40 ratio testing to Quest Diagnostics.

## 2 METHODS

### 2.1 Study design and clinical assessments

This cross-sectional study involved 250 participants who provided plasma specimens at the 1Florida Alzheimer’s Disease Research Center (ADRC cohort). The study was approved by the Mount Sinai Medical Center IRB, and all participants provided informed consent. Participants were between the ages of 50 and 95 years, had a minimum 6th grade reading comprehension level, and spoke English or Spanish as their primary language. Participants were excluded from the study if they had significant visual or auditory deficits or non-AD medical or psychiatric illness.

In the ADRC cohort, all participants underwent clinical assessments including an extensive medical, neurological, psychiatric, and neuropsychological evaluation. Assessments included the clinical dementia rating (CDR) and the mini-mental state exam (MMSE), and cognitive diagnoses were carried out using a previously reported algorithmic procedure.^17^ Participants were categorized as healthy controls (HC), having mild cognitive impairment (MCI), or AD.^18^ The cohort included 72 HC (ADRC-HC; 5 Aβ positive by PET [Aβ-PET+] and 67 negative [Aβ-PET-] by PET), 124 with MCI (ADRC-MCI; 42 Aβ-PET+ and 82 Aβ-PET-), and 54 with AD (ADRC-AD; all Aβ-PET+). Participant demographics are summarized in **Table 1** and **Table S1**.

**Table 1.** Alzheimer’s Disease Research Center (ADRC) cohort participant characteristics by amyloid positive emission tomography (Aβ PET) status.

| Characteristic | Aβ PET- [N = 149] | Aβ PET+ [N = 101] |
| --- | --- | --- |
| **Age (years) (*P* = 0.150)** |  |  |
| Mean (SD) [N] | 71.6 (7.5) [149] | 73.1 (8.5) [101] |
| **Sex (*P* = 0.794)** |  |  |
| % female [N] | 58.4% [87] | 60.4% [61] |
| **Ethnicity (*P* = 1.000)** |  |  |
| % Hispanic [N] | 59.1% [88] | 58.4% [59] |
| **Education (*P* = 0.234)** |  |  |
| Mean years (SD) | 15.6 (3.2) | 15.1 (3.2) |
| **MMSE (*P* < 0.001)** |  |  |
| % MMSE ≥ 25 [N] | 94.0% [140] | 57.4% [58] |
| Mean (SD) [N] | 28.4 (2.1) [149] | 23.6 (6.2) [101] |
| **CDR (*P* < 0.001)** |  |  |
| % CDR > 0 [N] | 66.4% [99] | 92.1% [93] |
| **Plasma Aβ42/40 (*P* < 0.001)** |  |  |
| Mean (SD) [N] | 0.173 (0.029) [149] | 0.141 (0.017) [101] |
| **Plasma Aβ42 (*P* < 0.001)** |  |  |
| Mean (SD) [N] | 68.3 (17.4) [149] | 58.4 (13.6) [101] |
| **Plasma Aβ40 (*P* = 0.079)** |  |  |
| Mean (SD) [N] | 394.5 (74.7) [149] | 411.5 (74.5) [101] |
| **ApoE4+, % (*P* < 0.001)** |  |  |
| % [N] | 20.8% [31] | 56.4% [57] |
| **<sup>18</sup>F-florbetaben SUVR (*P* < 0.001)** |  |  |
| Mean (SD) [N] | 1.020 (0.092) [93] | 1.469 (0.187) [71] |
| **<sup>18</sup>F-florbetapir SUVR (*P* < 0.001)** |  |  |
| Mean (SD) [N] | 1.042 (0.068) [26] | 1.455 (0.238) [15] |

Abbreviations: MMSE, Mini-mental state exam; CDR, Clinical dementia rating; SUVR, standardized uptake value ratio.

In addition, we performed a retrospective analysis of 6,192 consecutive plasma specimens submitted to Quest Diagnostics for Aβ42/40 ratio testing. This was a limited data set (https://privacyruleandresearch.nih.gov/pr_08.asp) with only patient sex and age information retained. Participant demographics are summarized in **Table 2**.

**Table 2.** Demographics for 6,192 patients.

| Age, years | N total, %<br>female | Aβ42 mean,<br>pg/mL (SD) | Aβ40 mean, pg/mL<br>(SD) | Aβ42/40 mean (SD) | Aβ42/40 <0.160, N, %<br>(indeterminant/high<br>risk of PET positivity) | Aβ42/40 ≥0.170, N, %<br>(likely PET negative) |
| --- | --- | --- | --- | --- | --- | --- |
| <40 | 83, 65% | 38.7 (8.3) | 207.3 (43.5) | 0.188 (0.021) | 3, 4% | 69, 83%, |
| 40 to 49 | 162, 62% | 38.2 (8.6) | 210.4 (42.4) | 0.182 (0.023) | 20, 12%, | 116, 72%, |
| 50 to 59 | 582, 63% | 40.5 (10.6) | 228.7 (57.2) | 0.178 (0.024) | 138, 24%, | 369, 63%, |
| 60 to 69 | 1,370, 58% | 40.6 (10.5) | 241.7 (58.5) | 0.169 (0.023) | 497, 36% | 631, 46%, |
| 70 to 79 | 2,451, 57% | 42.9 (11.9) | 264.7 (71.0) | 0.163 (0.022) | 1,157, 47% | 863, 35%, |
| 80 to 89 | 1,417, 54% | 45.5 (12.5) | 284.9 (77.8) | 0.161 (0.021) | 713, 50% | 436, 31%, |
| ≥ 90 | 127, 60% | 48.1 (10.7) | 304.5 (74.0) | 0.159 (0.019) | 68, 54%, | 32, 25%, |
| All | 6,192, 57% | 42.7 (11.7) | 259.4 (71.3) | 0.166 (0.023) | 2,596, 42%, | 2,516, 41%, |

### 2.2 Assessment of amyloid positivity from amyloid PET scans

Qualitative and quantitative analysis of amyloid positivity by visual reads and standardized uptake value ratio (SUVR) data have previously been described.^18^ All ADRC participants underwent an amyloid PET scan within 12 months of plasma collection. Of these individuals, 205 had SUVR computed using 2 different tracers: [F^18^]-florbetapir (n = 41, cutoff >1.11) and [F^18^]-florbetaben (n = 164, cutoff >1.40). SUVR values were transformed into a binary scale (Aβ-PET+ or Aβ-PET-) based on SUVR cutoffs. However, visual reads were used as the gold standard for amyloid positivity designations^18^ whenever there were conflicting data between SUVR cutoffs and visual reads. Amyloid PET statuses for the remaining 45 individuals were determined by visual reads alone.^18,19^

### 2.3 Plasma beta-amyloid and ApoE LC-MS/MS assays

Blood specimens were collected by venipuncture into 10 mL tubes containing EDTA as anticoagulant, kept on ice (<1 hour) until centrifugation at 1200 relative centrifuge force for 12 minutes at room temperature. Aliquots of plasma (0.5 mL) were transferred into polypropylene tubes and stored at -80 ⁰C until analysis. All plasma specimens were deidentified, and results were blinded during the analysis.

A detailed description of the plasma Aβ42/40 LC-MS/MS assay, as well as the LC-MS/MS plasma ApoE proteotype assay, will be reported elsewhere (Weber et al, submitted).^16^ Briefly, all mass spectrometry measurements were performed on a TSQ Altis Plus Triple Quadrupole MS (ThermoFisher Scientific, Waltham, MA) operated in multiple-reaction monitoring (MRM) mode. Assay precision (within-run and between-run), analytical measurement range (AMR), analytical sensitivity (limit-of-blank [LOB], limit-of-detection [LOD], and limit-of-quantification [LOQ]), interference testing, and stability were conducted according to CLSI guidelines.^20–24^

For this study, we determined average inter-assay (between-run) imprecision by taking the average of 4 quality control (QC) samples run in duplicate over 25 consecutive days (Weber et al. submitted; ranges were 163-619 pg/mL Aβ40, 40-188 pg/mL Aβ42, and 0.124-0.304 Aβ42/40; a non-physiological high QC value was excluded).^16^

Analytical robustness and the effects of reclassification bias were empirically determined by reanalyzing previously tested samples using a different operator and a different lot of reagents. A total of 196 plasma specimens were randomly selected and reanalyzed over the course of one month. Both freeze/thaw and storage stability were considered when selecting specimens for reanalysis. Values for Aβ40, Aβ42, and the Aβ42/40 ratio were compared as a percentile difference calculated between the mean values of the original results and the mean values of the retest results.

### 2.4 Statistical analysis

Two-sample t-tests were used to evaluate differences in baseline continuous measures across PET status. Differences in categorical variables were evaluated by Fisher’s exact test. Comparisons between groups with more than two outcomes were performed using one-way analysis of variance (ANOVA) for continuous variables and Fisher’s exact tests for categorical variables. Effect size for means were determined by eta-square measurements. Statistically significant results from ANOVA were followed by post-hoc analysis using Tukey multiple pairwise-comparisons between group means. The performance of the Aβ42/40 ratio on classification of amyloid PET status was evaluated by logistic regression and ROC curve analysis. Receiver operating characteristics (ROC) area under curve (AUC) 95% confidence intervals (CI), and comparisons between ROC curves were determined using the DeLong method.^25^ Optimal cutoffs for sensitivity and specificity from ROC analyses were determined by Youden’s J.^26^ Correlation of the Aβ42/40 ratio and PET SUVR were assessed by Spearman’s rho. Robustness simulations were based on the methodology described in Rabe, et al.^13^

To account for the potential underestimation of variability in the measure of the Aβ42/40 ratio in this study, 10,000 simulations of the Aβ42 and Aβ40 responses were generated with an added 10% CV (per Rabe, et al^13^) and 6% measured CV from a scaled standard normal distribution. For each simulated pair of markers with added noise, an Aβ42/40 ratio was calculated. The rate of reclassification around the 0.160 Aβ42/40 threshold from the original observed Aβ42/40 ratio to the simulated noise-added ratio was calculated. The average reclassification rate was estimated as the mean of the 10,000 simulated reclassification rates. A 95% confidence interval (CI) for the mean reclassification rate was estimated by the 2.5^th^ and 97.5^th^ percentiles of the simulated reclassification rates.

To estimate the effects of potential bias in the measure of Aβ42 and Aβ40 on the performance characteristics of the Aβ42/40 ratio, per Rabe, et al,^13^ we evaluated a 10% increase in Aβ42 response and a 10% decrease in the Aβ40 response for a total Aβ42/40 bias of 22% (1.1/0.1). The measured bias was similarly assessed. Performance characteristics were calculated for both the observed Aβ42/40 ratio and the biased Aβ42/40 ratio. Joint 95% CI were obtained for sensitivity/specificity and PPV/NPV on classification of PET status.^27^ All CI for performance characteristics were obtained by non-parametric bootstrap. All analyses were conducted using R (version 4.2.1).

## 3 RESULTS

### 3.1 ADRC cohort participant demographics: PET status and cognitive outcomes

Compared to ADRC Aβ-PET-individuals, Aβ-PET+ individuals were more likely to be *APOE4* carriers, have lower MMSE scores, and have a higher proportion with CDR scores greater than 0 (*P* < 0.001 for all, **Table 1**). No statistically significant differences (*P* >0.05) between the Aβ-PET- and the Aβ-PET+ group were observed in age or patient sex, percentage of individuals identifying as Hispanic, or years of education (**Table 1**).

When categorized by a cognitive diagnosis, from HC to MCI to AD (**Table S1**), we observed lower MMSE scores among MCI and AD patients compared to HC (*P* < 0.001, **Table S1**). In addition, the frequency of *APOE4* carriers was greater in the ADRC HC Aβ-PET+, MCI Aβ-PET+, and AD Aβ-PET+ groups compared to the HC Aβ-PET- and MCI Aβ-PET-group (*P* < 0.001). The ADRC-MCI Aβ-PET-group had a lower percentage of female participants relative to the other groups (*P* = 0.024) (**Table S1**). There were no statistically significant differences in PET status based on age, patient education, or percentage individuals identifying as Hispanic.

### 3.2 Performance and robustness assessment of plasma Aβ42/40 ratio for identifying amyloid PET status

Overall, for the 250 individuals with PET data (quantitative and qualitative reads), plasma Aβ42 concentrations and Aβ42/40 ratios were significantly lower (*P* < 0.001) in ADRC Aβ-PET+ individuals compared with Aβ-PET-individuals with no significant differences in Aβ40 concentrations (**Table 1**, **Figure 1A**). Using ROC analysis and the maximum of Youden’s J index, a plasma Aβ42/40 cutoff ratio of 0.160 had an AUC of 0.84 (95% CI = 0.79 to 0.89, **Figure 1B**) with 91% sensitivity, 76% specificity, and overall accuracy of 82% (**Table 3**). Based on a 40% prevalence of amyloid positivity in the ADRC cohort (40.4% observed), we found a positive predictive value (PPV) of 72% and a negative predictive value (NPV) of 93% at the 0.160 cutpoint (**Table 3**). Based on a 34% prevalence of amyloid positivity in the patients in the ADRC cohort with MCI, which may be more reflective of the target population, PPV decreased to 56% and NPV decreased to 90% at the 0.160 cutpoint (**Table 3**) when HC and AD patients were excluded.

**Figure 1:**
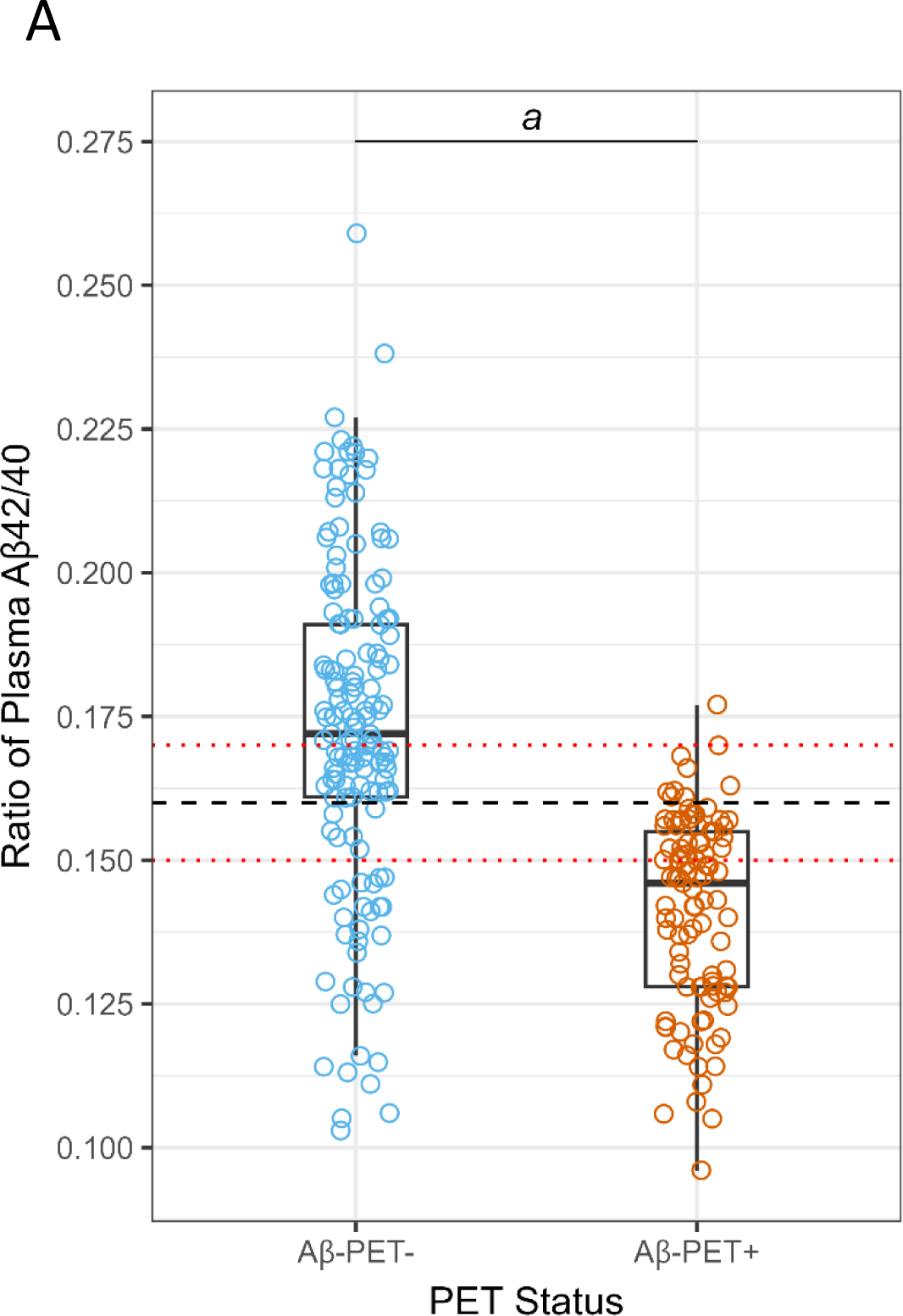
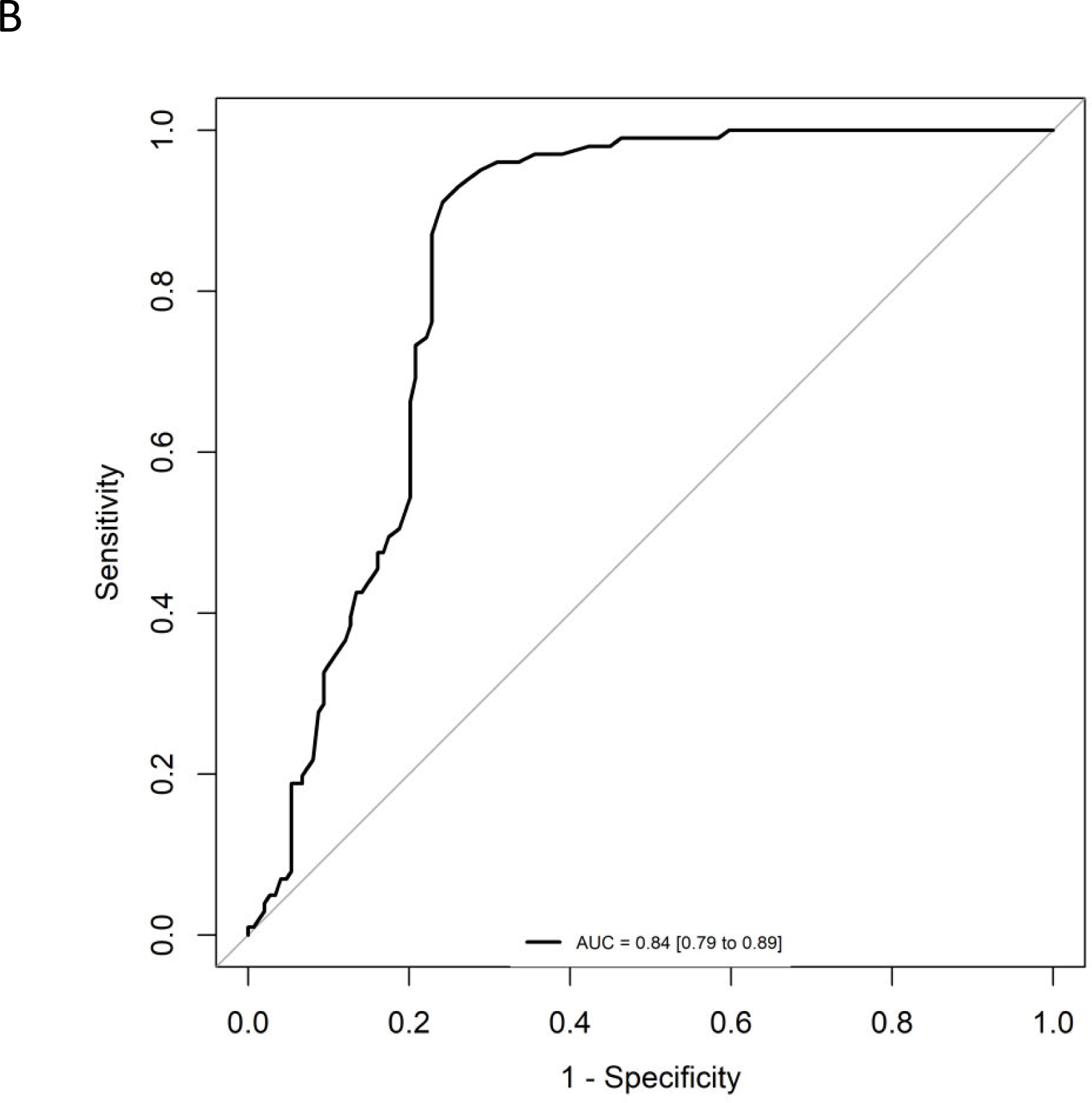
Correlation and diagnostic performance of the Aβ42/40 ratio and amyloid PET imaging. **A)** Plasma Aβ42/40 ratio compared with amyloid PET status (Aβ-PET- and Aβ-PET+). Black dashed line denotes optimal Aβ42/40 ratio cutoff of 0.160. Red dashed lines denote Aβ42/40 ratio indeterminant risk cutoffs (0.150 and 0.170); *a* = significant at *P* < 0.001; **B)** ROC-AUC of the plasma Aβ42/40 ratio for prediction of amyloid PET positivity. AUC, area under the curve; ROC, receiver operating characteristic.

**Table 3.** Performance characteristics for detecting positive emission tomography (PET) status in the Alzheimer’s Disease Research Center (ADRC) cohort at 0.160 (maximum of Youden’s J index) and 0.170 (likely PET-negative) cutpoints, and the effect of prevalence and bias (bold=performance based on measured prevalence of PET positivity in full ADRC cohort).

| Plasma Aβ42/40<br>Cutpoint | Prevalence<br>(%) | Bias<br>(%) | Sensitivity<br>(95%, CI) | Specificity<br>(95%, CI) | Accuracy<br>(95%, CI) | PPV<br>(95%, CI) | NPV<br>(95%, CI) |
| --- | --- | --- | --- | --- | --- | --- | --- |
| **0.160** | **40** | **none** | **0.911**<br>**(0.842-0.970)** | **0.758**<br>**(0.678-0.832)** | **0.820**<br>**(0.772-0.864)** | **0.719**<br>**(0.656-0.788)** | **0.926**<br>**(0.876-0.974)** |
| 0.160 | 34* | none | 0.857<br>(0.738-0.976) | 0.659<br>(0.537-0.768) | 0.726<br>(0.645-0.798) | 0.563<br>(0.478-0.661) | 0.900<br>(0.820-0.980) |
| 0.160 | 34* | 22<sup>†</sup> | 0.333<br>(0.167-0.500) | 0.854<br>(0.756-0.939) | 0.677<br>(0.605-0.742) | 0.538<br>(0.346-0.737) | 0.714<br>(0.663-0.771) |
| 0.160 | 34* | 2<sup>‡</sup> | 0.762<br>(0.619-0.905) | 0.671<br>(0.549-0.780) | 0.702<br>(0.621-0.782) | 0.542<br>(0.446-0.649) | 0.846<br>(0.762-0.932) |
| **0.170** | **40** | **none** | **0.990**<br>**(0.960-1.000)** | **0.537**<br>**(0.443-0.624)** | **0.720**<br>**(0.672-0.768)** | **0.592**<br>**(0.546-0.643)** | **0.988**<br>**(0.953-1.000)** |
| 0.170 | 34* | none | 0.976<br>(0.905-1.000) | 0.476<br>(0.354-0.598) | 0.645<br>(0.573-0.718) | 0.488<br>(0.433-0.554) | 0.975<br>(0.907-1.000) |
| 0.170 | 34* | 22<sup>†</sup> | 0.429<br>(0.262-0.595) | 0.829<br>(0.732-0.915) | 0.694<br>(0.621-0.766) | 0.563<br>(0.400-0.732) | 0.739<br>(0.681-0.802) |
| 0.170 | 34* | 2<sup>‡</sup> | 0.929<br>(0.833-1.000) | 0.537<br>(0.415-0.659) | 0.669<br>(0.589-0.742) | 0.506<br>(0.441-0.583) | 0.936<br>(0.851-1.000) |

\* Amyloid positivity by PET in the ADRC cohort—MCI group only
<sup>†</sup> +10% bias in Aβ42 and -10% bias in Aβ40 = 22% total bias per Rabe, et al.<sup>13</sup>
<sup>‡</sup> +2% total measured bias in the LC-MS/MS assay

The difference in means between the PET+ and PET-Aβ42/40 ratio was about 18% with substantial shift of Aβ PET+ to PET-status after applying 10% bias (22% total bias if Aβ42 and bias in Aβ40 shifted in opposite directions, **Figure 2A**) per Rabe, et al.^13^ This level of bias would substantially diminish the PPVs and NPVs of this assay (**Table 3**). However, we measured actual bias in this LC-MS/MS assay as 11.5% (95% CI = 9.5% to 13.6%) for Aβ42, 10.6% (95% CI = 9.0% to 12.6%) for Aβ40 (both in the same direction), and 0.7% (95% CI = -0.3% to 1.8%) for Aβ42/40 ratio. Accordingly, we used a higher estimate (upper 95% limit of the ratio CI rounded to 2%) to reflect worst-case effects of bias (**Figure 2A**). PPVs and NPVs are much less affected under these conditions (**Table 3**).

**Figure 2:**
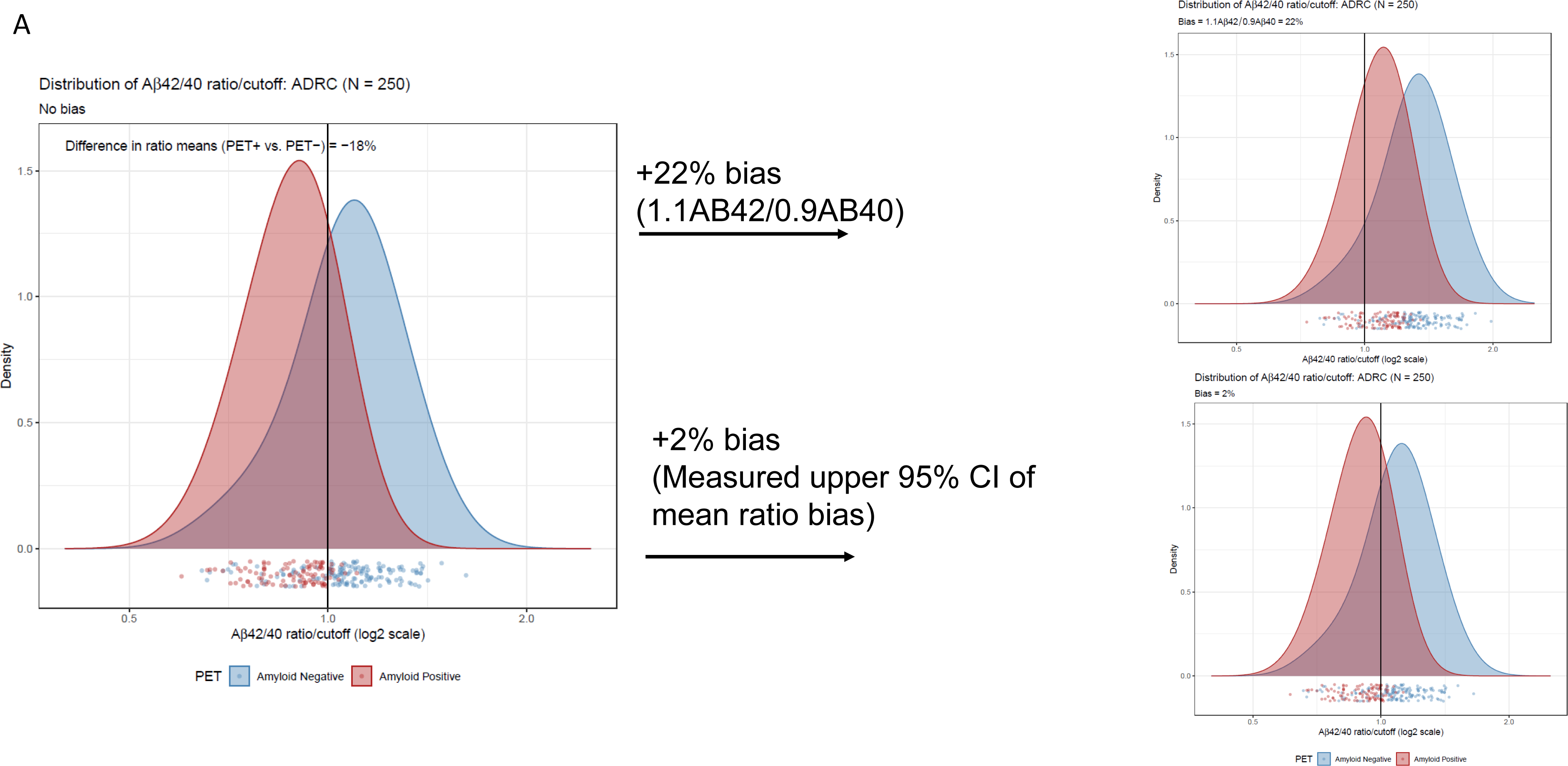
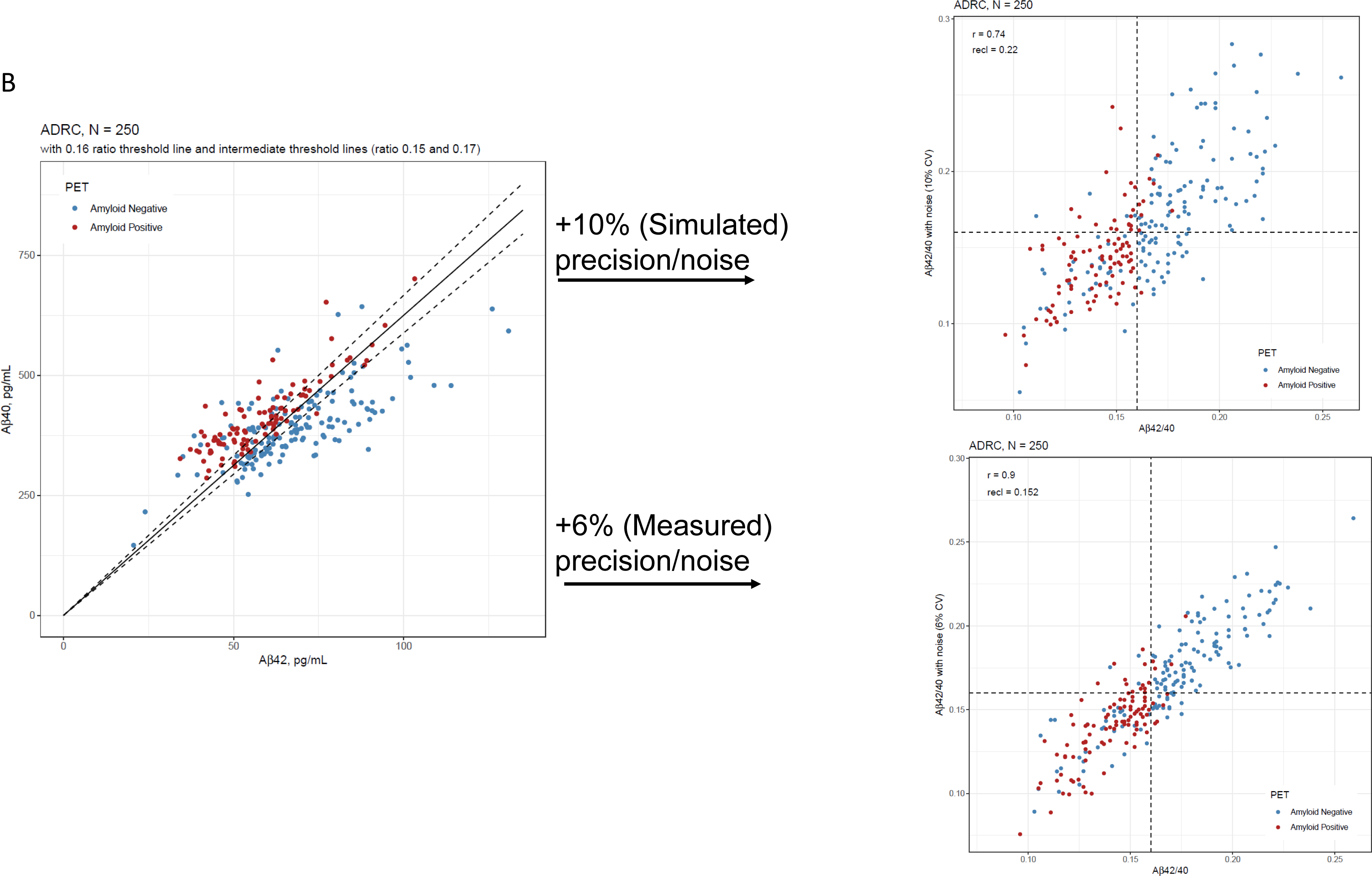
Robustness assessment of Aβ42/40 ratio for predicting amyloid PET imaging results. **A)** Densities of plasma Aβ42/40 ratio by LC-MS/MS in ADRC cohort with and without 10% bias (22% total, upper plot) and 2% measured bias (lower plot) added. **B)** Scatterplots of Aβ42 and Aβ40 to illustrate proximity to cutoffs defining indeterminate risk and scatterplots of Aβ42/40 ratio with and without 10% CV added noise (upper plot) and 6% added CV (measured) noise (lower plot).

Assuming an estimated imprecision of 10% per Rabe, et al^13^ we simulated a reclassification rate of 22% (95% CI= 18% to 27%) in the ADRC cohort at a cutpoint of 0.160 (**Figure 2B upper scatterplot**). However, actual average inter-assay imprecision is closer to 6% (Weber et al, submitted).^16^ Using this measured precision, the reclassification rate by simulation is 15% (95% CI= 11% to 19%) (**Figure 2B lower scatter plot**).

Based on 6% assay imprecision, we propose cutpoints of 0.150 and 0.170 as defining indeterminate risk of amyloid PET positivity, constituting 30% of the ADRC cohort (**Figure 2B**, **Table 4**). The latter cutpoint has an NPV of 99% in the ADRC cohort for ruling out amyloid positivity (98% in the target population with MCI, 94% when adding measured bias, **Table 3**). Large differences were observed within the ranges defined by indeterminate risk in the ADRC cohort including 4- to 5-fold higher PET positivity and ApoE4 phenotype (*APOE4* genotype) and 9-fold higher AD diagnosis, in the 0.150-0.159 range vs the 0.160-0.169 range (**Table 4**).

**Table 4.** Proposed plasma Aβ42/40 ranges for clinical decision making. Percentage of individuals falling within a range unless otherwise specified. Real world projections of positive emission tomography (PET) positivity are based on percent PET positivity in the Alzheimer’s Disease Research Center (ADRC) cohort.

| **Plasma Aβ42/40 Range** | **Designation** | **ADRC, N (% of cohort)** | **PET positivity, N (%)** | **MCI, N (%)** | **AD, N (%)** | **ApoE4, N (%)** | **Real world, N (% of cohort)** | **Real world, projections of PET positivity, N** | **Potential next evaluation and management considerations** |
| --- | --- | --- | --- | --- | --- | --- | --- | --- | --- |
| <0.150 | Positive (low ratio) | 91 (36.4) | 61 (67.0) | 50 (54.9) | 34 (37.4) | 43 (47.3) | 1468 (23.7) | 984 | Follow up for AD workup (high risk) |
| 0.150-0.159 | Indeterminant (lower ratio) | 37 (14.8) | 31 (83.8) | 14 (37.8) | 18 (48.7) | 25 (67.6) | 1128 (18.2) | 945 | Follow up for AD workup (indeterminant risk) |
| 0.160-0.169 | Indeterminant (higher ratio) | 38 (15.2) | 7 (18.4) | 19 (50.0) | 2 (5.3) | 7 (18.4) | 1080 (17.4) | 199 | Follow up for AD workup (low risk) |
| ≥0.170 | Negative (high ratio) | 84 (33.6) | 2 (2.4) | 41 (48.8) | 0 (0.0) | 13 (15.5) | 2516 (40.6) | 60 | Follow up for non-AD etiology |
| All | All | 250 | 101 (40.4) | 124 (49.6) | 54 (21.6) | 88 (35.2) | 6192 | 2188 |  |

For the 205 individuals for whom we had quantitative PET data, 5 [F^18^]-florbetapir positive results (range 1.11 to 1.15) were reclassified as negative and 25 [F^18^]-florbetaben negative results (range 1.09 to 1.39) were reclassified as positive based on visual reads. After reclassification, the concordance of the Aβ42/40 ratio increased from 80% to 88% for [F^18^]-florbetapir and from 74% to 83% [F18]-florbetaben. Overall concordance between Aβ42/40 ratio and SUVR data changed from 75% before to 84% after reclassification based on visual reads.

Both tracers showed similar results when plotted against the Aβ42/40 ratio (**Figures 3A and 3B**). We observed a significant inverse relationship between the Aβ42/40 ratio and quantitative SUVR values for [F^18^]-florbetapir PET (Spearman’s rho of -0.58 (95% CI = -0.76 to -0.32, *P* <0.001), **Figure 3A**) and [F^18^]-florbetaben PET (Spearman’s rho of -0.54 (95% CI = -0.65 to -0.42, *P* <0.001), **Figure 3B**).

**Figure 3:**
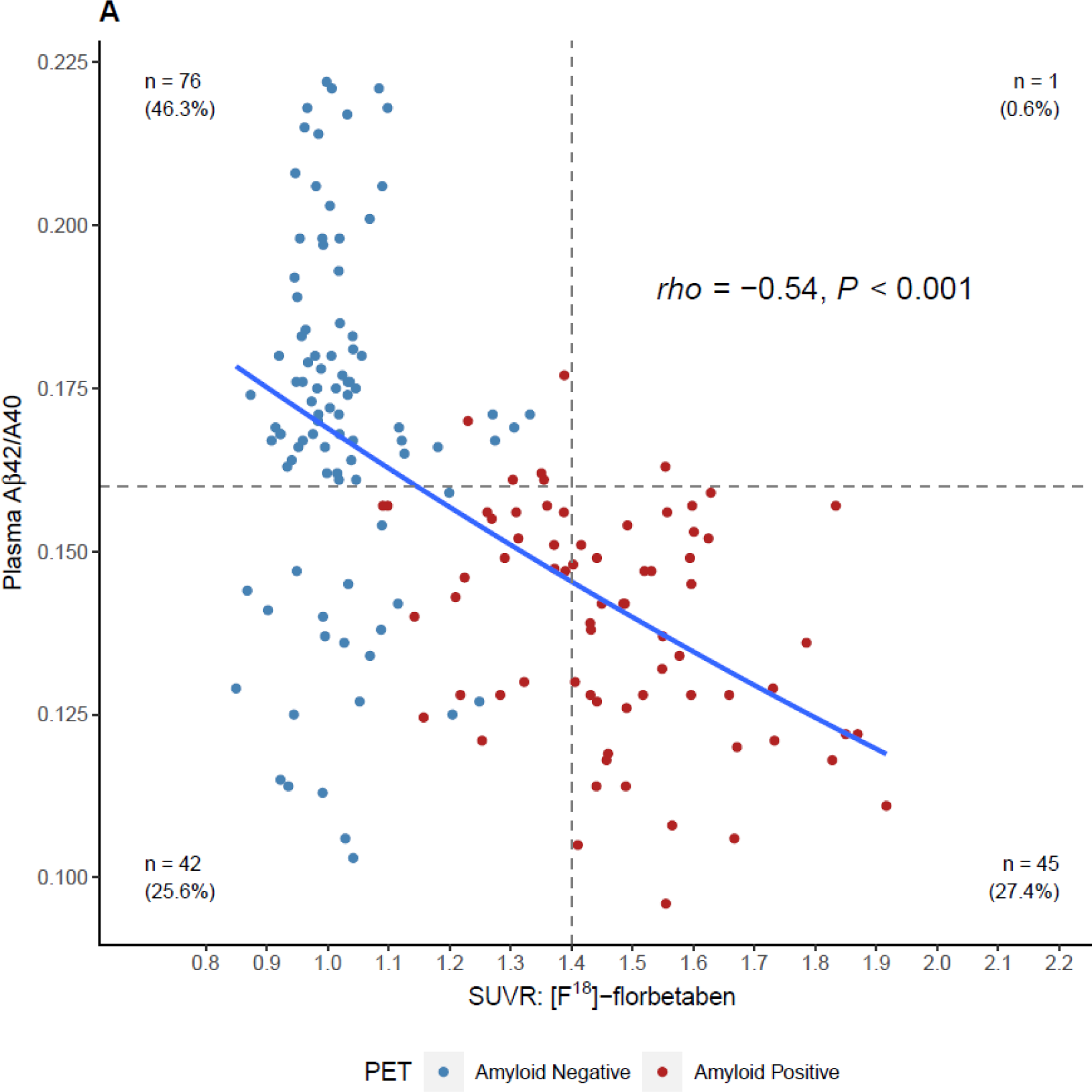
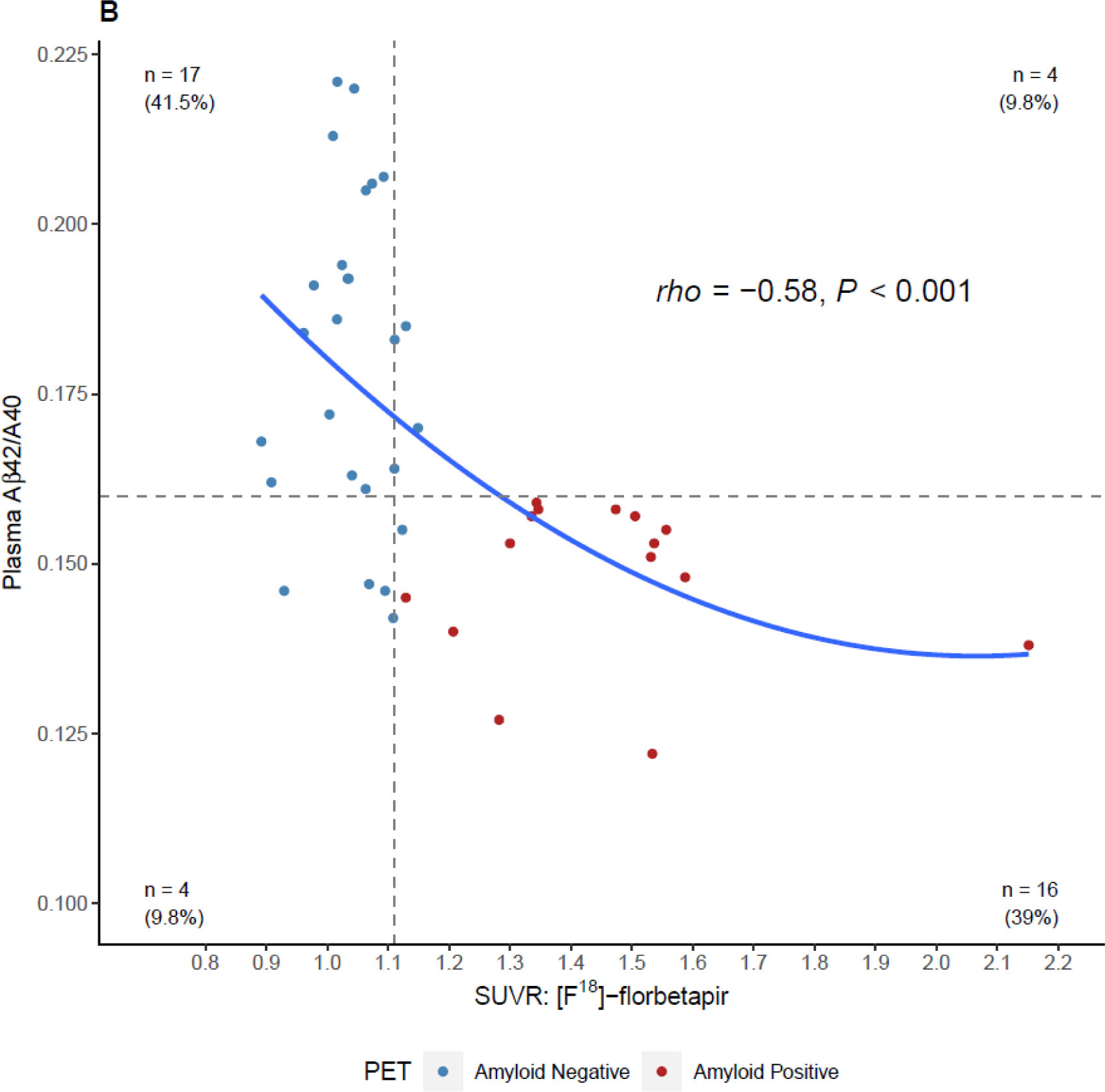
Four-quadrant plot illustrating the relationship between the plasma Aβ42/40 ratio and **A)** [F^18^]-florbetaben SUVR values and, B) [F^18^]-florbetapir SUVR values. Horizontal dashed line = optimal plasma Aβ42/40 ratio cutoff (0.160). Vertical dashed lines = optimized SUVR cutoff values for each tracer. Color coding for amyloid positivity and negativity is based SUVR cutoffs or gold-standard visuals reads. SUVR, standardized uptake value ratio.

### 3.3 Plasma Aβ42/40 ratio and clinical specimens

A significant inverse relationship between age and Aβ42/40 ratio was observed for the 6,192 clinical specimens (Spearman’s rho = -0.25, 95% CI = -0.27 to -0.23, *P* < 0.001; **Figure 4a**, **Table 2**), contrasting with the increases of plasma Aβ42 and Aβ40 concentrations with age (**Figure 4b and c**, **Table 2**). Indeterminate results defined by cutpoints of 0.150 and 0.170 define 2,208 (35.7%) almost evenly split between individuals with ratios from 0.150 to 0.159 and individuals with ratios from 0.160 to 0.169, but with the former group potentially representing a much higher percentage of PET-positivity based on the ADRC cohort (**Table 4**). Using a 0.170 cutpoint to indicate the likelihood of PET negativity, we identify 2,516 individuals for whom a PET scan/CSF would be potentially unnecessary. As expected, among these individuals, the percentage who are likely PET negative decreases with age from <40 (83%) to ≥ 90 years (25%) (**Table 2**).

**Figure 4:**
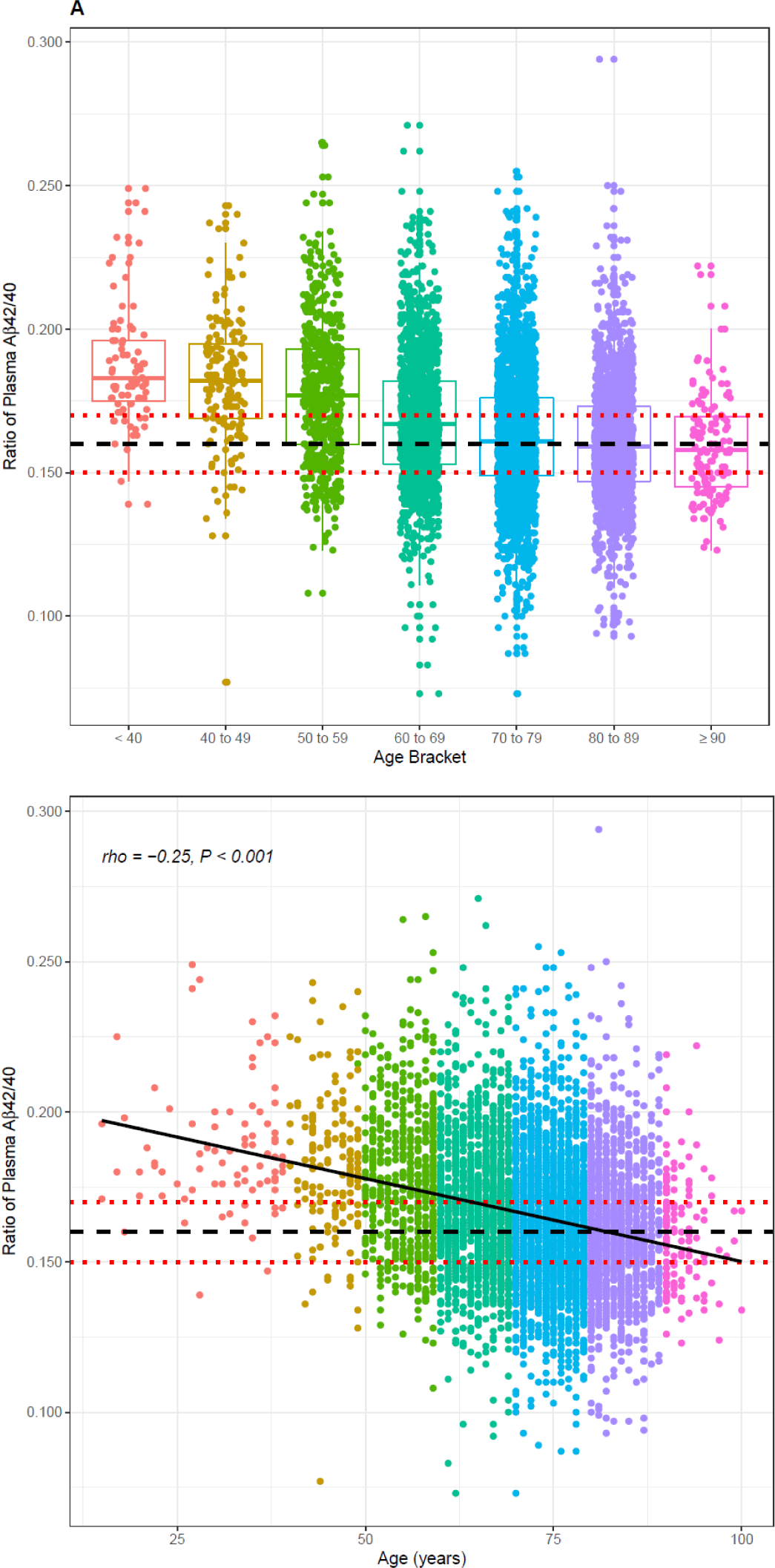
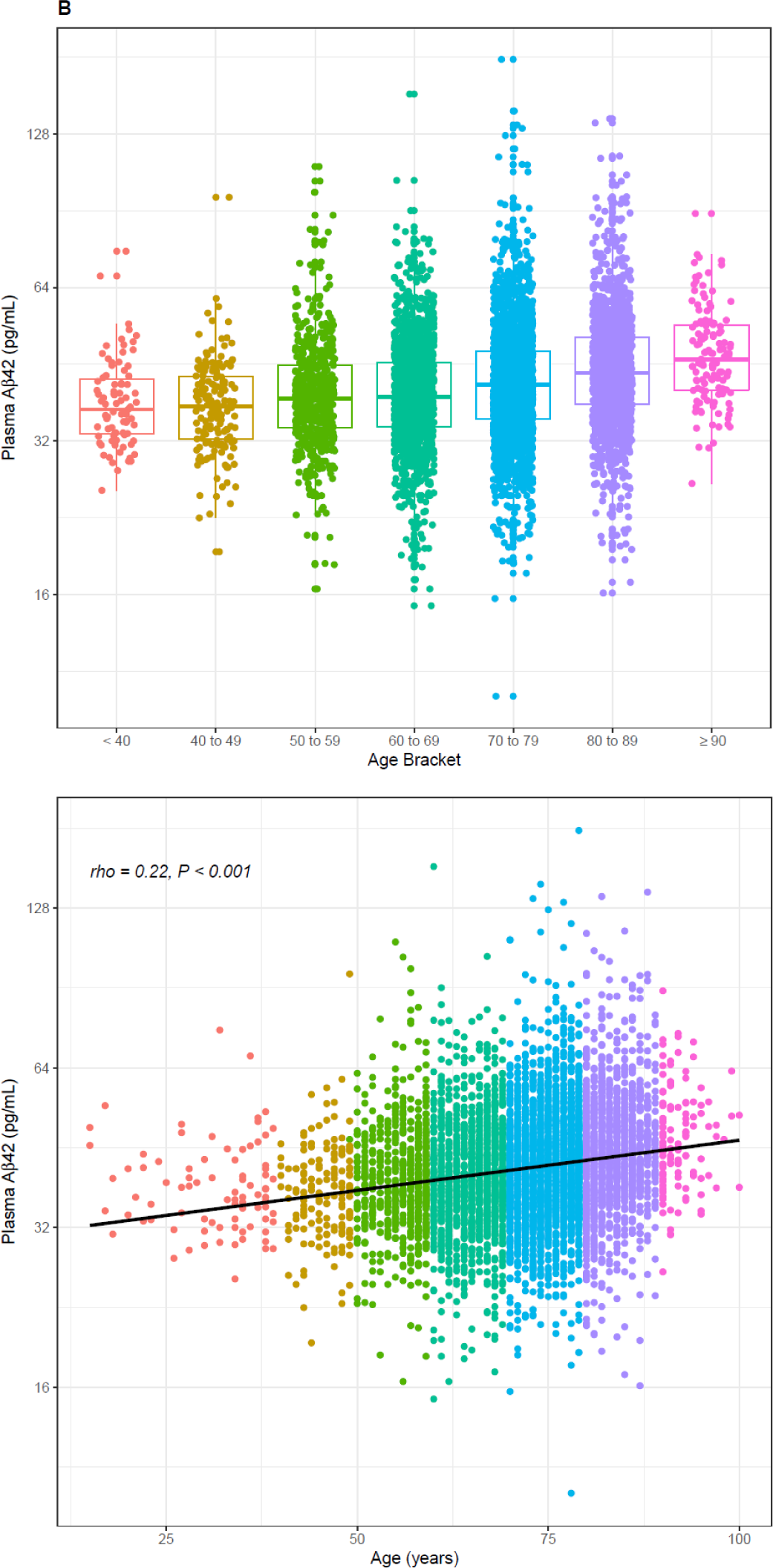
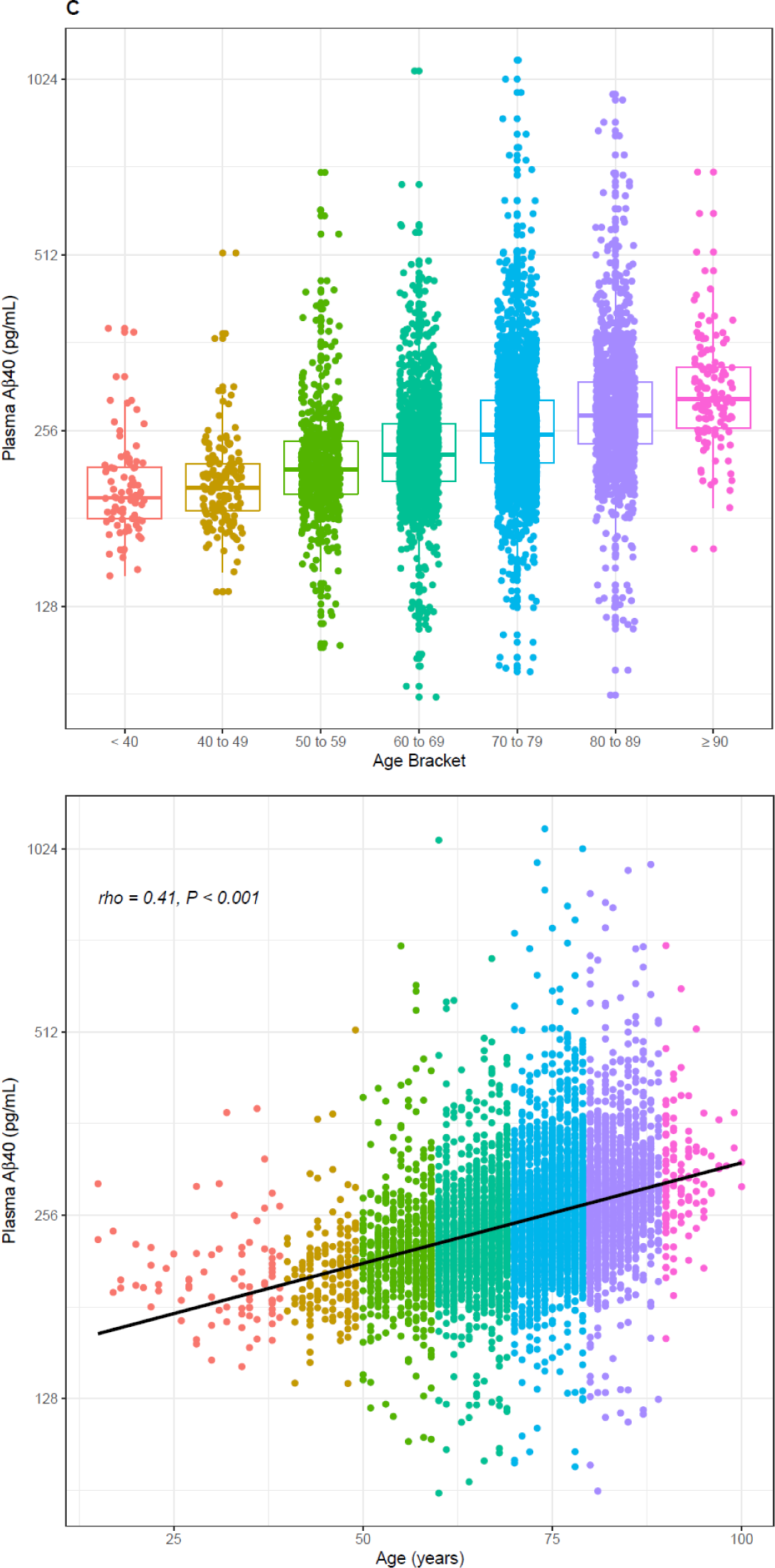
Distribution (upper) and scatterplots (lower, *P* <.001 for all) by age of; **A)** Aβ42/40 ratio, Spearman’s rho = -0.25, 95%CI, -0.27 to -0.22. **B)** Aβ42 concentrations, Spearman’s rho = 0.22, 95%CI 0.20 to 0.25; and **C)** Aβ40 concentrations, Spearman’s rho = 0.41, 95%CI 0.39 to 0.43. Black dashed line denotes Aβ42/40 ratio cutoff of 0.160. Red dashed lines denote Aβ42/40 ratio indeterminant risk cutoffs (0.150 and 0.170).

## 4 DISCUSSION

Our data support the use of plasma Aβ42/40 ratio by LC-MS/MS to help predict a low likelihood of PET-positivity in AD assessment. In the ADRC group, with an Aβ42/40 ratio cutpoint of 0.170, we observed a 99% NPV. Only 2 of 84 (2.4%) patients with a higher Aβ42/40 ratio had PET positivity. Although previous studies have cast doubt on the clinical utility of plasma Aβ42/40 assays based on misclassification potential, the only detailed analysis was provided for the Elecsys Aβ42 and Aβ40 immunoassays. LC-MS/MS assays, although acknowledged to have better discriminatory performance as assessed by AUROC analysis, were said to have similar issues in terms of narrow analytical ranges and fold changes between Aβ-PET+ and PET-individuals yielding insufficient robustness for clinical decision making.^13^

We followed the example of Rabe et al^13^ by applying these tools to the results for the ADRC cohort obtained using our new LC-MS/MS assay. The ADRC cohort has a moderate prevalence of amyloid PET positivity (40%), within the range of 17% to 60% previously reported for cohorts studied by LC-MS/MS.^28,29^

In keeping with previous studies, we found that the plasma Aβ42/40 ratio was significantly lower in individuals that were Aβ-PET+ compared to those that were Aβ-PET-. Performance characteristics were comparable to other LC-MS/MS assays; however, we found higher fold changes (18%) between means of PET+ and PET-individuals (**Figure 2A**) than were previously reported (6% to 14%).^13^

Applying a 10% bias (in opposite directions, total 22% per Rabe, et al^13^) negatively impacted assay sensitivity, specificity, accuracy, NPV, and PPV (**Table 3**). However, applying a measured mean Aβ42/40 ratio bias (0.7%, worst case 2%) had a much lesser effect (**Figure 2A**), despite mean bias for each analyte being close in magnitude to the simulations for Aβ42 (∼12%) and Aβ40 (∼11%). Under real-word conditions, bias for each analyte drifted in the *same direction,* unlike the simulations, effectively canceling out its effect on the ratio. We note that our measured bias is similar to the 1% “drift” Aβ42/40 ratio values reported by Hu et al in their high-throughput MS assay.^28^

Sources of bias that affect accurate determination of the Aβ42/40 ratio are those that negatively impact one analyte over the other, effectively skewing the ratio over time. For example, specimen storage at room temperature can result in differential loss of Aβ42 relative to Aβ40,^30^ suggesting that Aβ42 is more susceptible to proteolytic cleavage, aggregation, or absorptive loss under these conditions. However, proper handling and storage mitigates these risks in the clinical laboratory. Immunoassays may be more susceptible to matrix effects that affect one analyte over the other compared with LC-MS/MS assays^31^ wherein analytical losses and suppression are corrected for using isotopically labeled internal standards. Errors in calibrator concentrations based on inaccurate supplier-provided peptide content can also cause bias, which we avoid by employing independent quantitative amino acid analysis of peptide calibrators.^32^ All these factors are impediments to standardization of these assays and may explain the high variability of Aβ42/40 ratio across assays and laboratories.^31^ Interestingly, the 0.160 Aβ42/40 ratio cutoff optimized for maximum sensitivity and specificity for detecting PET positivity is identical to the optimized cutoff for differentiating HCs from individuals with AD, both in the current plasma study (Figure S1) and in CSF.^33^ This suggests that the proportion of Aβ40 and Aβ42 is similar in CSF and plasma.

Applying a simulated 10% imprecision per Rabe, et al^13^ yielded a relatively high reclassification rate for individuals in our ADRC cohort (22%), similar to their immunoassay and the BioFINDER cohort (26%). However, measured imprecision was closer to 6%, yielding a reclassification rate of 15%. Although this still represents a substantial group of individuals, it should not unduly affect the clinical utility of this assay for identifying those with a low likelihood of PET-positivity.

Our clinical data set was obtained from 6,192 consecutively run Aβ42/40 ratio specimens, presumably submitted by healthcare providers to help understand the cause of memory decline and dementia, and potentially help diagnose AD. The trends we observed in our data set, namely that plasma Aβ42/40 decreased as plasma Aβ42 and Aβ40 concentrations increased with age, generally aligned with those recently reported in China for ∼200 specimens obtained from apparently healthy, older (50-89 years) individuals.^34^

Based on cutpoints defining high, low, and indeterminant risk in the ADRC cohort, we suggest that plasma Aβ42/40 ratio results can help guide clinical decision making (**Table 4**). Notably, the high cutpoint of 0.170 almost rules out PET-positivity with an NPV of almost 99% (acknowledging potential variations caused by prevalence and bias in **Table 3**). Projecting percentages of PET-positivity at this cutpoint in the ADRC cohort onto the 6,192 clinical specimens, the assay could potentially negate the need for PET testing in 2,516 patients (**Table 4**). Assuming a PET scan cost of $5,000,^35^ this approach saves $12,580,000 after subtracting the cost of 6,192 LC-MS/MS analyses (at current list pricing) to identify these patients and 60 PET scans for the projected 2.4% patients with false negative results who would eventually need a PET scan. This represents a total savings of $9.2 million or about $1,485 per patient. Exceptions to this clinical decision making may include some patients with autosomal dominant AD and both amyloid precursor protein and presenilin-1 mutations who may have an elevated Aβ42/40 ratio.^36^

Our results support those from studies using comparable IP-LC-MS/MS assays^37–41^ for Aβ42/40; the ROC-AUC (0.84) is similar to those reported previously for discriminating PET positivity.^38,39,41^ However, the inclusion of age and the number of *APOE4* alleles did not improve classification accuracy based on ROC analysis, despite many studies that demonstrated significant improvements when age and *APOE4* status were added to their models.^15,28,39,41^

Age did not improve the model predictor for PET positivity because the Aβ PET+ and Aβ PET-individuals were age matched. However, age-dependent trends for the Aβ42/40 ratio in our clinic data set, as well as the reference range studies conducted by Chen et al,^34^ suggest that NPV could be influenced by age, especially for individuals under 60 years of age who tend to have higher Aβ42/40 ratios.

*APOE4* status may not have improved the model predictor because of the demographics of our cohort. Previous studies included predominantly White and/or Asian populations.^15,28,39,41^ Notably, the association between the *APOE4* allele and AD risk has been reported to be lower in Hispanic and Black non-Hispanic individuals than in White non-Hispanic individuals ^42,43^. Our ADRC cohort had roughly 3-times the number of Hispanic individuals (59.1% of the Aβ PET+ and 58.4% of the Aβ PET-individuals) compared with similar studies that showed a significant improvement in AUC when *APOE4* status was included. ^39^ Our findings are a cautionary note that algorithms incorporating Aβ42/40 and *APOE4* allele status may not be generally applicable for all races and ethnicities.

### Limitations

We did not perform a detailed comparison with other biomarkers in the current study; instead, we focused on the utility of the plasma Aβ42/40 ratio biomarker in isolation. However, in a separate investigation, we show that early AD-related pathological changes in the Aβ42/40 biomarker were associated with quantifiable changes in brain microstructure and connectivity in Aβ-PET negative patients preceding deviations in other plasma biomarkers including t-tau, p-tau, neurofilament light chain (NfL), and glial fibrillary acidic protein (GFAP), and cortical atrophy (DeSimone et al, submitted).^44^

We did not examine adding an additional plasma biomarker such as p-tau or glial fibrillary acidic protein (GFAP). Some studies have combined plasma biomarkers to enhance performance in predicting clinical outcome. Addition of plasma p-tau with plasma Aβ42/40 might enhance the diagnosis in patients with MCI, because plasma p-tau levels typically increase with disease progression.^45^ The combination of biomarkers could be used in patients with specific clinical or demographic characteristics to further help define individual outcomes.^46^ The combination of Aβ42/40 and p-tau may be better able to predict cognitive decline, but performance of the individual assays used would also play a role in how well the combination aids with this prediction.^47^ In addition, the combination of Aβ42/40 and GFAP enhanced the likelihood of becoming p-tau positive, suggesting that this biomarker might distinguish AD patients with progressive features.^48^

Other study limitations include a relatively small sample size for individuals with PET data and insufficient racial diversity (predominantly Hispanic) that may pose a potential bias to our results, especially when it comes to the effect of the *APOE4* allele on AD status. The lack of longitudinal plasma specimens from individuals that either converted from being Aβ-PET- to Aβ-PET+, or individuals that transitioned from being cognitively normal to MCI and AD limit our current understanding of the prognostic utility of the Aβ42/40 ratio in monitoring disease progression and identifying patients who may benefit from disease-modifying therapeutics.

## 5 CONCLUSIONS

Our IP-LC-MS/MS assay accurately identified individuals with positive amyloid PET and differentiated individuals diagnosed with AD from age-matched HC. These findings support the use of this blood-based assay for assessing presence of AD pathology in individuals with cognitive impairment and can help reduce PET evaluations in patients with low likelihood of AD pathology, allowing for more efficient allocation of limited imaging resources.

## Data Availability

All data produced in the present study are available upon reasonable request to the authors

## ACKNOWLEDGEMTS

We acknowledge helpful comments from Drs Lee Hilborne, Harvey Kaufman, Matt Stroh, and Andrew Hellman. Funding for this work was provided by National Institutes of Health (NIH) Center Core Grant P30AG066506 (TEG) and NIH Research Grants R01NS052318 and R01NS075012 (DEV).

## CONFLICT OF INTEREST STATEMENT

Darren M. Weber, Steven W. Taylor, Robert J. Lagier, Jueun C. Kim, Scott M. Goldman, Nigel J. Clarke, and Michael K. Racke are employees of Quest Diagnostics. All other authors have nothing to disclose.

## CONSENT STATEMENT

All study participants provided informed consent prior to undergoing study procedures.

## Notes

### Clinical Trial

This study is of a new plasma LC-MS/MS assay for the Aβ42/Aβ40 ratio

### Author Declarations

The study was approved by the Mount Sinai Medical Center IRB, and all participants provided informed consent.

