## Supplemental material for "Clinical utility of plasma Aβ42/40 ratio by LC-MS/MS in Alzheimer’s disease assessment"

**Table S1.** ADRC participant characteristics by cognitive diagnosis and amyloid PET status

| Characteristic | Healthy Controls (HC) | | | Mild Cognitive Impairment (MCI) | | | Alzheimer's Disease (AD) | PET status* | | | Diagnosis† | | |
|---|---|---|---|---|---|---|---|---|---|---|---|---|---|
| | Combined [N = 72] | HC Aβ- [N = 67] | HC Aβ+ [N = 5] | Combined MCI [N = 124] | MCI Aβ- [N=82] | MCI Aβ+ [N=42] | AD Aβ+ [N=54] | F | *P*-value‡ | Eta-Square | F | *P*-value‡ | Eta-Square |
| **Demographics** | | | | | | | | | | | | | |
| **Age** | | | | | | | | | | | | | |
| Mean (SD) | 70.3 (7.2) | 70.4 (7.3) | 69.2 (4.9) | 73.1 (7.5) | 72.6 (7.5) | 74.1 (7.4) | 72.7 (9.4) | 1.81 | 0.128 | 0.03 | 3.10 | 0.047 | 0.02 |
| **Sex** | | | | | | | | | | | | | |
| % female [N] | 72.2% [52] | 71.6% [48] | 80.0% [4] | 50.8% [63] | 47.6% [39] | 57.1% [24] | 61.1% [33] | NA | 0.039 | NA | NA | 0.013 | NA |
| **Education** | | | | | | | | | | | | | |
| Mean years (SD) | 16.3 (2.9) | 16.2 (2.9) | 16.8 (1.6) | 15.2 (3.2) | 15.1 (3.3) | 15.3 (3.0) | 14.8 (3.4) | 2.06 | 0.086 | 0.03 | 4.00 | 0.019 | 0.03 |
| **Ethnicity** | | | | | | | | | | | | | |
| % Hispanic | 59.7% [43] | 62.7% [42] | 20% [1] | 57.3% [71] | 56.1% [46] | 58.1% [25] | 62.3% [33] | NA | 0.459 | NA | NA | 0.888 | NA |
| **MMSE** | | | | | | | | | | | | | |
| Mean (SD) | 29.1 (1.4) | 29.1 (1.4) | 29.4 (0.9) | 27.7 (2.4) | 27.8 (2.4) | 27.4 (2.5) | 20.1 (6.3) | 60.98 | < 0.001 | 0.50 | 122.63 | < 0.001 | 0.50 |
| **CDR** | | | | | | | | | | | | | |
| % CDR > 0 [N] | 33.3% [24] | 35.8% [24] | 0% [0] | 91.9% [114] | 91.5% [75] | 92.9% [39] | 100% [54] | NA | < 0.001 | NA | NA | < 0.001 | NA |
| ***APOE* ε4** | | | | | | | | | | | | | |
| 1 or 2 ε4 alleles (% [N]) | 23.6% [17] | 20.9% [14] | 60% [3] | 32.3% [40] | 20.7% [17] | 55.8% [23] | 57.4% [31] | 11.17 | < 0.001 | 0.15 | NA | < 0.001 | NA |
| **Plasma Biomarkers** | | | | | | | | | | | | | |
| **Aβ42, pg/mL** | | | | | | | | | | | | | |
| Mean (SD) | 68.9 (15.2) | 69.8 (15.3) | 56.5 (6.4) | 64.4 (17.4) | 67.0 (18.9) | 59.3 (12.7) | 57.9 (14.9) | 6.07 | < 0.001 | 0.09 | 7.08 | 0.001 | 0.05 |
| **Aβ40, pg/mL** | | | | | | | | | | | | | |
| Mean (SD) | 387.5 (58.7) | 388.8 (60.2) | 370.4 (30.3) | 405.9 (77.6) | 399.2 (84.8) | 418.9 (60.2) | 409.5 (85.9) | 1.44 | 0.220 | 0.02 | 1.78 | 0.171 | 0.01 |
| **Aβ42/40** | | | | | | | | | | | | | |
| Mean (SD) | 0.177 (0.026) | 0.179 (0.025) | 0.152 (0.008) | 0.159 (0.030) | 0.168 (0.031) | 0.141 (0.019) | 0.141 (0.015) | 27.26 | < 0.001 | 0.31 | 30.76 | < 0.001 | 0.20 |
| **Amyloid PET Imaging Tracer** | | | | | | | | | | | | | |
| **$^{18}$F-florbetaben SUVR** | | | | | | | | | | | | | |
| Mean (SD) | 1.007 (0.068) | 1.007 (0.068) | NA | 1.190 (0.248) | 1.031 (0.109) | 1.450 (0.183) | 1.482 (0.191) | 135.90 | < 0.001 | 0.72 | 60.62 | < 0.001 | 0.43 |
| **$^{18}$F-florbetapir SUVR** | | | | | | | | | | | | | |
| Mean (SD) | 1.089 (0.140) | 1.057 (0.069) | 1.531 (NA) | 1.154 (0.211) | 1.023 (0.064) | 1.415 (0.138) | 1.474 (0.312) | 16.78 | < 0.001 | 0.65 | 9.16 | < 0.001 | 0.33 |

Aβ, beta-amyloid; CDR, clinical dementia rating; MMSE, mini-mental state examination; NA, not applicable; PET, positron emission tomography; SD, standard deviation; SUVR, standard uptake value ratio

* ANOVA analysis performed on HC Aβ-/+, MCI Aβ-/+, and AD Aβ+ groups only

† ANOVA analysis performed on Combined HC, MCI, and AD groups

‡ For continuous variables global *P*-value from ANOVA F-test. Otherwise, Fisher's exact test

**Figure S1.** Correlation and diagnostic performance of the Aβ42/40 ratio with cognitive diagnosis for the ADRC cohort. **A)** Plasma Aβ42/40 ratio compared with healthy controls (ADRC-HC), mild cognitive impairment (ADRC-MCI), and Alzheimer's disease (ADRC-AD). Amyloid PET negative (Aβ-PET-) individuals are illustrated with blue circles, and amyloid PET positive (Aβ-PET+) individuals are illustrated with orange circles; **B)** ROC-AUC of the plasma Aβ42/40 ratio for ADRC-HC vs. ADRC-AD. *a* = Significant at $P < 0.001$

A

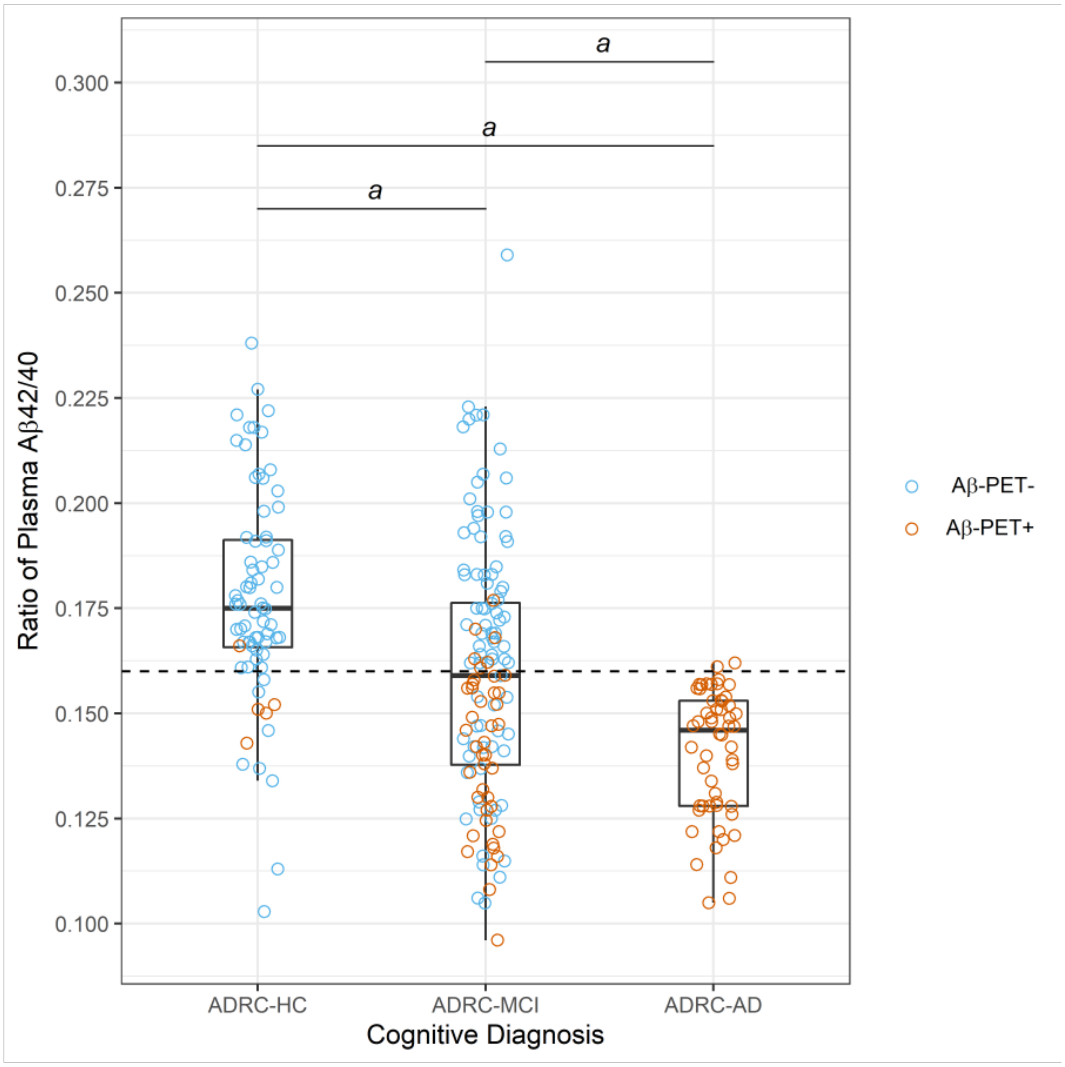

B

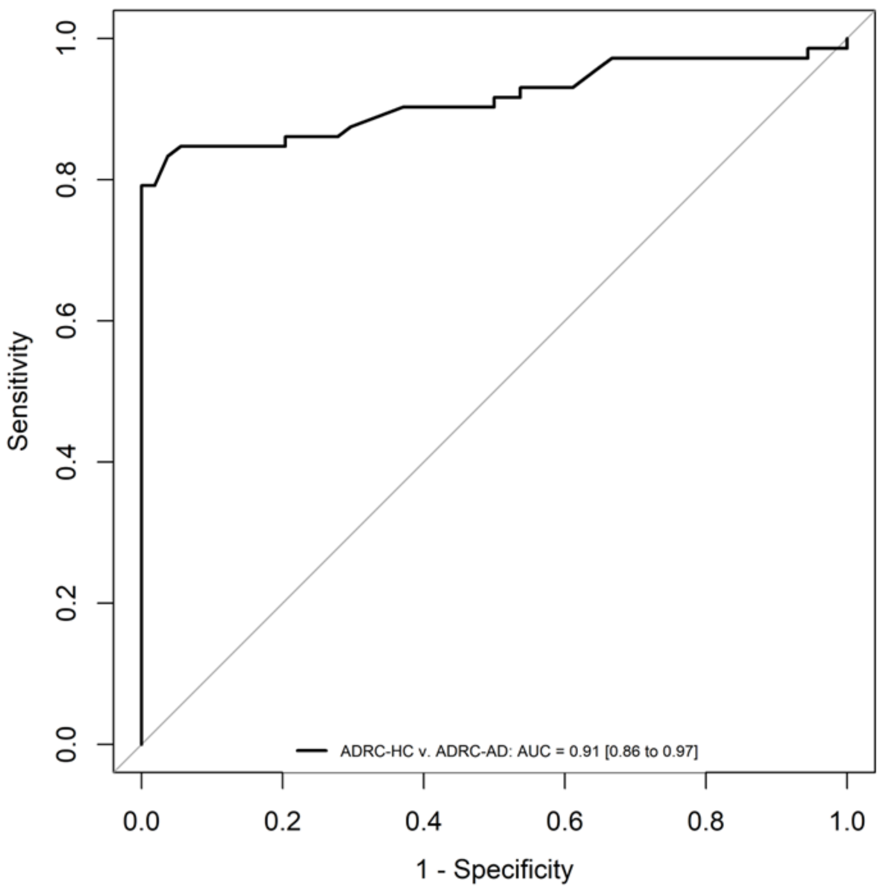

Sensitivity
1.0
0.8
0.6
0.4
0.2
0.0
ADRC-HC v. ADRC-AD: AUC = 0.91 [0.86 to 0.97]
0.0
0.2
0.4
0.6
0.8
1.0
1 - Specificity
